# Similarities and Differences in the Late-Onset GM2 Gangliosidoses: Tay-Sachs and Sandhoff Diseases

**DOI:** 10.1101/2025.08.05.25333048

**Authors:** Connor J. Lewis, Leila Shirvan, Jean M. Johnston, Catherine Groden, John Yang, Andrea Ashton, Jessica Chong, Mark Moran, Hera Akmal, Selby I. Chipman, Cris Zampieri, Jordan Wickstrom, Jesse Matsubara, Frances Gavelli, Tanya Lehky, Katharine E. Alter, Camilo Toro, Cynthia J. Tifft

**Author notes:** Corresponding Author: Cynthia J Tifft. These authors contributed equally to this work (Connor J. Lewis, Leila Shirvan).

## Abstract

The two predominating subtypes of late-onset GM2 gangliosidosis are late-onset Tay-Sachs (LOTS) and late-onset Sandhoff disease (LOSD). Due to shared deficiencies of ß-hexosamindase A and significant clinical overlap, the two diseases have been considered indistinguishable. However, a growing body of evidence supports the notion of several distinctions between the two diseases. In this study, we highlight these distinctions through the cross-sectional evaluation of 27 late-onset GM2 gangliosidosis participants. Twenty-one participants with LOTS and 6 with LOSD were included in this study. We performed physical examinations alongside assessments for gait, balance, muscle strength, ataxia, nerve conduction velocities, and analyzed brain magnetic resonance imaging. Lower limb weakness (95% in LOTS, 100% in LOSD) and later development of upper limb weakness (90% in LOTS, 83% in LOSD) was highly prevalent in both cohorts. Accompanying gait disturbances, balance issues, and dysmetria (as assessed by the brief ataxia rating scale [BARS]) were also prevalent in both cohorts. Strength testing for the quadriceps and hamstrings demonstrated weakness in both cohorts, primarily impacting extensor muscles. Supratentorial gray and white matter volumes in both cohorts were similar to normative data. In contrast, BARS scores for dysarthria and oculomotor dysfunction were present and heterogenous in LOTS participants and absent in LOSD participants. 24% of LOTS participants and none of the LOSD participants had a history of neuropsychiatric symptoms. Cerebellar volume including lobules V and VI were lower in LOTS compared to LOSD and normative data. However, length dependent sensory neuropathy was present in all LOSD participants but absent in LOTS participants. Dysfunction of the posterior cerebellum (lobules VI, VII, and IX) has been shown to cause cerebellar cognitive affective syndrome (CCAS), that includes cognitive and behavioral disturbances. Furthermore, cerebellar dysfunction of lobules V and VI has been linked to dysarthric speech, and dysfunction of the posterior cerebellum has been linked to oculomotor symptoms. The finding of low cerebellar lobule volumes in LOTS, suggests the distinctive features of the LOTS phenotype are related to cerebellar dysfunction. However, the sensory symptoms unique to LOSD remains a mystery. The molecular and biochemical basis for the dichotomy between the LOTS and LOSD phenotypes requires further investigation.

## Introduction

The GM2 gangliosidoses are rare autosomal recessive lysosomal storage disorders caused by the accumulation of GM2 ganglioside due to the absence or near absence of ß-hexosamindase A (EC 3.1.2.52), the heterodimeric enzyme that degrades glycosphingolipid GM2 in healthy tissue of unaffected individuals. Pathological accumulation of GM2 ganglioside unleashes a cascade of events leading to progressive neurodegeneration.^1,2^ Tay-Sachs and Sandhoff diseases are the predominate forms of GM2 gangliosidosis with a prevalence of 1 in 200,000-320,000 and 1:500,000-1,500,000 individuals, respectively.^3,4,5^ An additional ultrarare form caused by biallelic variants in *GM2A*, encodes the GM2 the activator protein also required for GM2 degradation.^6^ The prevalence of Tay-Sachs disease is higher among the Ashkenazi Jewish and non-Jewish French Canadians from southeastern Quebec due to founder effects.^5,7,8,9^ Tay-Sachs disease (OMIM #272800) is caused by biallelic pathogenic mutations in *HEXA*, encoding the alpha subunit of ß-hexosaminidase A, while Sandhoff disease (OMIM #268800) is caused by biallelic pathogenic mutations in *HEXB*, which encodes the beta subunit of ß-hexosaminidase A.^2,10,11^

Both Tay-Sachs and Sandhoff diseases are a continuum of disease severity classified into infantile, juvenile, and late-onset (adult) forms based on symptom onset and roughly correlates with residual ß-hexosaminidase A activity.^12,13^ The infantile forms of these diseases are the most severe with symptom onset before six months and death by five years of age.^14,15^ The juvenile form of these diseases is less severe with symptom onset between 2 and 6 years of age and death within the second decade. The adult or late-onset form of Tay-Sachs or Sandhoff diseases are the most heterogeneous, with symptom onset during adolescence or early adulthood and life expectancy slightly reduced compared to unaffected individuals.^16,17,18^ There is no targeted treatment for the GM2 gangliosidoses, however trials of gene therapy, and the modified amino acid *N*-acetyl-L-leucine (NALL) have been recently reported.^19,20^

Participants with late-onset GM2 present with neuromuscular weakness, cerebellar dysfunction, and/or psychiatric symptoms.^17,18^ There is substantial heterogeneity in presenting symptoms and disease progression of participants, even among siblings sharing the same mutations and genetic background.^21,22,23^ Neurodegeneration is inexorably progressive over decades with worsening weakness, incoordination, dysarthria, and, in the later stages of the disease, cognitive dysfunction.^18,24,25^ Late-onset Tay-Sachs (LOTS) and late-onset Sandhoff disease (LOSD) have long been considered clinically indistinguishable. However, some phenotypic features of LOTS and LOSD appear more prevalent in one form over the other. For example, psychosis is more characteristic of LOTS while early sensory symptoms seem to be more pronounced in LOSD.^17^ Pure cerebellar dysfunction and dysarthria may be more severe in LOTS.^24,25^ However, weakness and ambulation difficulties are common to both.^18^ Strength testing on physical examination shows earlier involvement in the lower limbs, more specifically in the hip flexor and knee extensor muscles with later progression to the upper limbs, involving primarily the triceps. Electrophysiological studies indicate neurogenic weakness consistent of an anterior horn neuronopathy, a shared phenotype of LOTS and LOSD.^10^

Magnetic resonance imaging (MRI) studies have demonstrated numerous findings at different stages of the disease,^10,26^ however cerebellar atrophy with associated enlargement of the 4^th^ ventricle is reported in both LOTS and LOSD.^27,28^ One study, including only participants with LOTS, analyzed specific lobules of the cerebellum and suggested a link between atrophy in cerebellar lobule VI to ataxia, dysarthria, and tremors in LOTS participants.^29^ Cerebellar atrophy may be more common in LOTS compared to LOSD.^28^ Due to the slow progression of the disease, capturing clinically reliable metrics for disease progression has been difficult. A study evaluating potential clinical outcomes, suggested the nine-hole peg test (9HP), the Friedreich’s Ataxia rating scale (FARS), and the assessment of intelligibility of dysarthric speech (AIDS) were promising tests for assessing disease progression.^30^

In this study, we conducted cross-sectional evaluations in a cohort of LOTS (*n* = 21) and LOSD (*n* = 6) participants enrolled in a natural history study. Specifically, we collected data from instrumented gait and balance analysis, lower limb (knee flexor and extensor) muscle strength dynamometry, nerve conduction velocities (NCVs) and needle electromyography (EMG), speech intelligibility, and brain MRI anatomical, volumetric, and advanced imaging analysis. This comprehensive dataset was intended to capture the nature and spectrum of functional impairment in late-onset GM2 disease, explore phenotypic differences between LOTS and LOSD, and correlate aspects of the clinical history and presenting symptoms to additional disease phenotypes.

## Methods

### Participants

Participants diagnosed with LOTS or LOSD were enrolled at the National Institutes of Health (NIH) Clinical Center under the “Natural History of Glycosphingolipid and Glycoprotein Disorders” (ClinicalTrials.gov identifier: NCT00029965).^31^ The NIH Institutional Review Board approved this protocol (02-HG-0107). Informed consent was completed with all participants prior to participation, and all research was completed in accordance with the Declaration of Helsinki. Participants were evaluated between May 2012 and July 2025 at the National Institutes of Health Clinical Center in Bethesda, Maryland. Participants underwent a variety of clinical evaluations including a physical examination, gait analysis, balance testing, and dynamometry strength testing, electrodiagnostic investigation, speech, and magnetic resonance imaging (MRI). Although the order of testing (functional testing, strength, balance, and gait) varied between participants, a rest period of at least 20 minutes was provided between testing sessions to allow participants to recover from possible fatigue. For participants with repeated evaluations, the visit with the most complete data in terms of testing was selected. Demographics and medical histories were reported using a questionnaire modified from Bley et al.^32^ Some participants were assisted by a family member due to difficulty writing or inability to recall details of their disease progression. Diagnosis was confirmed by enzyme analysis in peripheral blood leukocytes and/or mutation analysis of *HEXA* and *HEXB* genes performed in CLIA-certified laboratories. 21 LOTS and 6 LOSD participants were included and further information on the participants are provided in Table 1. Some participants did not complete all the tests described in this study because of personal choice or safety concerns.

**Table 1.**
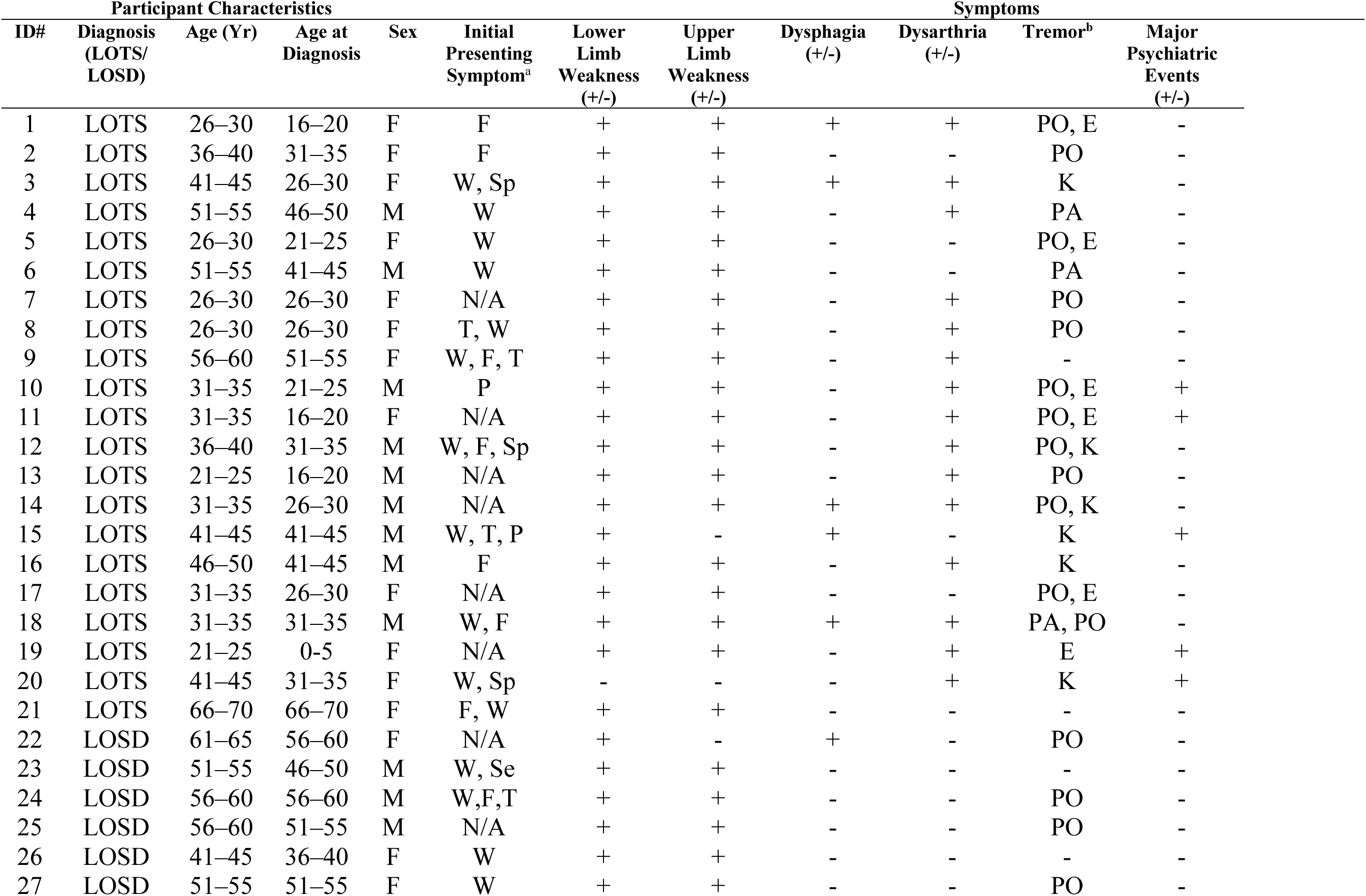
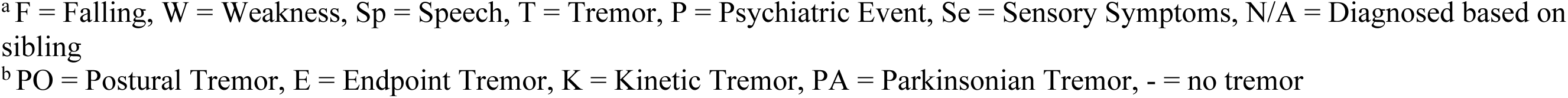
Summary of Participant Characteristics.

| Participant Characteristics |  |  |  |  |  | Symptoms |  |  |  |  |  |
| --- | --- | --- | --- | --- | --- | --- | --- | --- | --- | --- | --- |
| ID# | Diagnosis (LOTS/LOSD) | Age (Yr) | Age at Diagnosis | Sex | Initial Presenting Symptom<sup>a</sup> | Lower Limb Weakness (+/-) | Upper Limb Weakness (+/-) | Dysphagia (+/-) | Dysarthria (+/-) | Tremor<sup>b</sup> | Major Psychiatric Events (+/-) |
| 1 | LOTS | 26–30 | 16–20 | F | F | + | + | + | + | PO, E | - |
| 2 | LOTS | 36–40 | 31–35 | F | F | + | + | - | - | PO | - |
| 3 | LOTS | 41–45 | 26–30 | F | W, Sp | + | + | + | + | K | - |
| 4 | LOTS | 51–55 | 46–50 | M | W | + | + | - | + | PA | - |
| 5 | LOTS | 26–30 | 21–25 | F | W | + | + | - | - | PO, E | - |
| 6 | LOTS | 51–55 | 41–45 | M | W | + | + | - | - | PA | - |
| 7 | LOTS | 26–30 | 26–30 | F | N/A | + | + | - | + | PO | - |
| 8 | LOTS | 26–30 | 26–30 | F | T, W | + | + | - | + | PO | - |
| 9 | LOTS | 56–60 | 51–55 | F | W, F, T | + | + | - | + | - | - |
| 10 | LOTS | 31–35 | 21–25 | M | P | + | + | - | + | PO, E | + |
| 11 | LOTS | 31–35 | 16–20 | F | N/A | + | + | - | + | PO, E | + |
| 12 | LOTS | 36–40 | 31–35 | M | W, F, Sp | + | + | - | + | PO, K | - |
| 13 | LOTS | 21–25 | 16–20 | M | N/A | + | + | - | + | PO | - |
| 14 | LOTS | 31–35 | 26–30 | M | N/A | + | + | + | + | PO, K | - |
| 15 | LOTS | 41–45 | 41–45 | M | W, T, P | + | - | + | - | K | + |
| 16 | LOTS | 46–50 | 41–45 | M | F | + | + | - | + | K | - |
| 17 | LOTS | 31–35 | 26–30 | F | N/A | + | + | - | - | PO, E | - |
| 18 | LOTS | 31–35 | 31–35 | M | W, F | + | + | + | + | PA, PO | - |
| 19 | LOTS | 21–25 | 0–5 | F | N/A | + | + | - | + | E | + |
| 20 | LOTS | 41–45 | 31–35 | F | W, Sp | - | - | - | + | K | + |
| 21 | LOTS | 66–70 | 66–70 | F | F, W | + | + | - | - | - | - |
| 22 | LOSD | 61–65 | 56–60 | F | N/A | + | - | + | - | PO | - |
| 23 | LOSD | 51–55 | 46–50 | M | W, Se | + | + | - | - | - | - |
| 24 | LOSD | 56–60 | 56–60 | M | W, F, T | + | + | - | - | PO | - |
| 25 | LOSD | 56–60 | 51–55 | M | N/A | + | + | - | - | PO | - |
| 26 | LOSD | 41–45 | 36–40 | F | W | + | + | - | - | - | - |
| 27 | LOSD | 51–55 | 51–55 | F | W | + | + | - | - | PO | - |

### Functional Testing

Functional testing was performed by the same research nurse (JMJ) for all participants. Functional testing included the Rolyan 9-Hole Peg (9HP) test, The Timed Get Up and Go Test (TGUG), and Trail Making Test Part A and B (TMT-A/B). The 9HP test was performed twice bilaterally, and the times (in seconds) from the dominant hand were averaged and reported. The TGUG test was performed with the participant sitting in a neutral position in an armchair, and the time (in seconds) required to stand up, walk 10 feet, and return to the chair were recorded in seconds. TMT-A and TMT-B were performed using the participant’s dominant hand.

### Ataxia

Ataxia was evaluated using the brief ataxia rating scale (BARS) by a qualified neurologist (CT).^33^ The complete BARS is scored out of 30, with five domains including gait (8 points), knee-tibia test (4 points for each side), finger-to-nose test (4 points for each side), dysarthria (4 points), and oculomotor abnormalities (2 points). A blank BARS scoring sheet is included in the Supplementary Material (Supplemental Figure 1). When the examiner could not commit between two scores on a BARS item, an intermediate score was provided.

### Gait Analysis

Gait was evaluated with a 6-minute walk test. Participants walked in a quiet, well-lit hospital hallway at their preferred pace for six minutes. They were instructed to walk close to the railing on one side of the corridor and perform turns when they reached the end of a 25-meter walking path. A physical therapist walked behind the participant for safety. To avoid falls due to fatigue, participants were reminded that they were not required to complete the task and could stop at any time. A 4.8-meter instrumented walkway (GAITRite; CIR Systems Inc., Sparta, NJ) was placed within the walking path.^34,35^ The walkway allowed for capturing the participant’s steady state gait (avoiding the acceleration and deceleration gait phases at the ends of the walking path). The footprints for each pass on the instrumented walkway were analyzed for gait speed (centimeters per second), step length (centimeters), step width (centimeters), and double support time (seconds). All passes were averaged as well as right and left sides for analysis of each parameter. Age and sex matched controls were extracted from McKay et al.^36^

### Balance Testing

Study participants performed the Modified Clinical Test of Sensory Interaction on Balance (mCTSIB) as described in Antoniadou et al.,^37^ on a NeuroCom Balance SMART Equitest® System (previously Natus Medical Inc., Seattle, OR, USA). Participants were asked to stand quietly, barefoot, and completely still for 10 seconds. The trial was performed 3 times under each of the following conditions and in the following order: standing on firm surface with eyes open, standing on firm surface with eyes closed, standing on foam surface with eyes open and standing on foam surface eyes closed. A given condition was not tested if the participants was unable to stand barefoot on the surface or if they or the therapist felt the test was unsafe. The sway velocity of the center of gravity is the variable measured in this test. An average sway velocity of 3 trials for each condition was used in the data analysis. Higher sway velocity scores indicate poorer postural control. Scores reaching a maximum of 6 degrees/second indicated a fall. For participants who were unable to complete all trials due to safety concerns, a value of 6 degrees per second (worst possible score) was assigned. Age and sex matched controls were obtained directly from NeuroCom Balance Systems Customer Support.

### Quadriceps and Hamstring Testing

A Biodex System 3 (Shirley, NY) was used to measure isometric and isokinetic knee extensor (quadriceps) and flexor (hamstring) strength. Participants were seated in the Biodex chair with the knee joint aligned along the dynamometer axis. The leg was secured with a strap across the thigh and the dynamometer arm secured near the malleoli. Participants completed 3 trials of isometric knee flexion and extension for 5 seconds separated by 15 second rests to allow for recovery. Knee flexion trials were measured starting from the participant’s maximum extension, and knee extension trials starting from the participant’s maximum flexion.

Participants completed 3 self-paced isokinetic flexion and extension trials at 60 degrees/second and a 30 second rest period between flexion and extension trials. Flexion was tested from participants’ end range knee extension while seated in the Biodex to the maximum flexion angle that could be achieved before the dynamometer arm contacted the Biodex chair. Extension was tested from the knee flexed, with the dynamometer arm against the Biodex chair, to participants’ end range knee extension. The maximum force in Newtons (N) during each trial was recorded and converted to Torque (Nm). The average of the three trials was calculated and normalized in relation to the participant’s weight in kilograms (Nm/kg). Control data were obtained from healthy volunteers recruited by the Rehabilitation Medicine Department of the Clinical Center, NIH.

### Sensory Examination and Electrodiagnostic Testing

Nerve conduction studies were performed using standard methodology on a Natus Viking Select or EDX machine (Natus, Middleton, WI). In general, the median and sural sensory nerves and the fibular and median motor nerves were tested, though the specific selection of nerves depended on clinical status. Needle EMG was performed using a concentric needle and filter settings of 2k–10K with spontaneous and motor unit potential activity recorded according to standard methodology. Choice of muscle for needle EMG studies was dependent on the distribution of weakness. Active neurogenic changes in this study were marked by the presence of fibrillation potentials and positive sharp waves (fibs/psw) with or without the presence of complex repetitive discharges (CRDs). Chronic neurogenic changes in this study were marked by the presence of long duration and/or high amplitude motor unit potentials with possible polyphasia. Generally, decreased recruitment was observed in the presence of neurogenic changes. Parameters were compared to department-based normative data rather than explicit controls.

### MRI

As described in Lewis et al.,^27^ T1-weighted magnetic resonance imaging (MRI) from 23 GM2 Natural History participants were acquired on a Phillips Achieva 3T system with an 8-channel SENSE head coil. 18 LOTS participants and 5 LOSD had MRI imaging at the cross-sectional visit selected. Volumetric analysis of MRI data was performed using Freesurfer’s (v7.4.1) *recon-all* reconstruction pipeline to calculate volumes of the gray matter, white matter, cerebellum, 4^th^ ventricles, and total intracranial volume (ICV). and thalamus.^27,38–46^ One thousand thirty-three neurotypical controls (NC) were also analyzed from OpenNeuro and included participants from the following studies: “The National Institute of Mental Health (NIMH) Intramural Healthy Volunteer Dataset” (*n* = 154),^47,48^ “Neurocognitive aging data release with behavioral, structural, and multi-echo functional MRI measures” (*n* = 298),^49,50^ “Paingen_placebo” (*n* = 395),^51,52^ and “AgeRisk” (*n* = 186)^53,54^ consisting of participants between 18 and 89 years old. T1-weighted MRI were also analyzed at the cerebellar lobule level as described in Lewis et al. to calculate volumes of lobules V and VI.^55^

### Statistical Analysis

Due to the small sample size, particularly in the LOSD cohort, data was descriptive in nature. Ordinal BARS scores were presented as medians and inner quartile ranges. Otherwise, when appropriate, data are reported as means (± SD). Data analysis and graph generation were performed in GraphPad Prism software for macOS, version 10.1.0 (GraphPad Software).

## Results

### Participants and Clinical Description

Twenty-one participants with LOTS (from 16 families, 12 females) and 6 participants with LOSD (from 4 families, 3 females) were evaluated under this protocol. Demographic data are presented in Table 1. At the time of this cross-sectional evaluation, 20 LOTS participants (95%) and all 6 LOSD participants had lower limb weakness, 19 LOTS participants (90%) and 5 LOSD participants (83%) had upper limb weakness from physical examination. Cerebellar dysfunction was present in 20 LOTS participants (95%) and 3 LOSD participants (50%). Dysphagia was present in 5 LOTS participants (24%) and 1 LOSD participant (17%), 15 LOTS participants (71%) and no LOSD had dysarthria, 5 LOTS participants (24%) and no LOSD participants had a history of a major psychiatric event. No LOTS participants and all 6 LOSD participants had evidence of sensory neuropathy and dysautonomia.

### Symptom Onset

Participant reported symptom onset in the LOTS and LOSD cohorts occurred at an average age of 13.8 ± 7.0 years and 18.2 ± 11.2 years, respectively (Figure 1A). The large range of age at diagnosis in LOTS participants (0-67 years) is due in part to one participant who was diagnosed at a very young age (Table 1). The average age of diagnosis was 33.0 ± 12.7 years in the LOTS participants and 51.5 ± 6.6 years in the LOSD participants (Figure 1B), indicating there is an average diagnostic delay of 19.2 years and 33.3 years, from the onset of symptoms, respectively. On average, gait impairment (Figure 1D) was first noticed earlier in the LOTS participants (22.0 ± 7.6 years) than in the LOSD participants (36.4 ± 4.7 years); however, five LOTS participants and 1 LOSD participant indicated they were “not sure” when they first experienced impaired gait. LOTS participants (24.8 ± 8.8 years) also required the earlier use of handrails to climb stairs compared to LOSD participants (29.5 ± 12.2 years); however, two LOTS participants (aged 33 and 41) did not require handrails at the time of this evaluation (Figure 1C).

**Figure 1.**
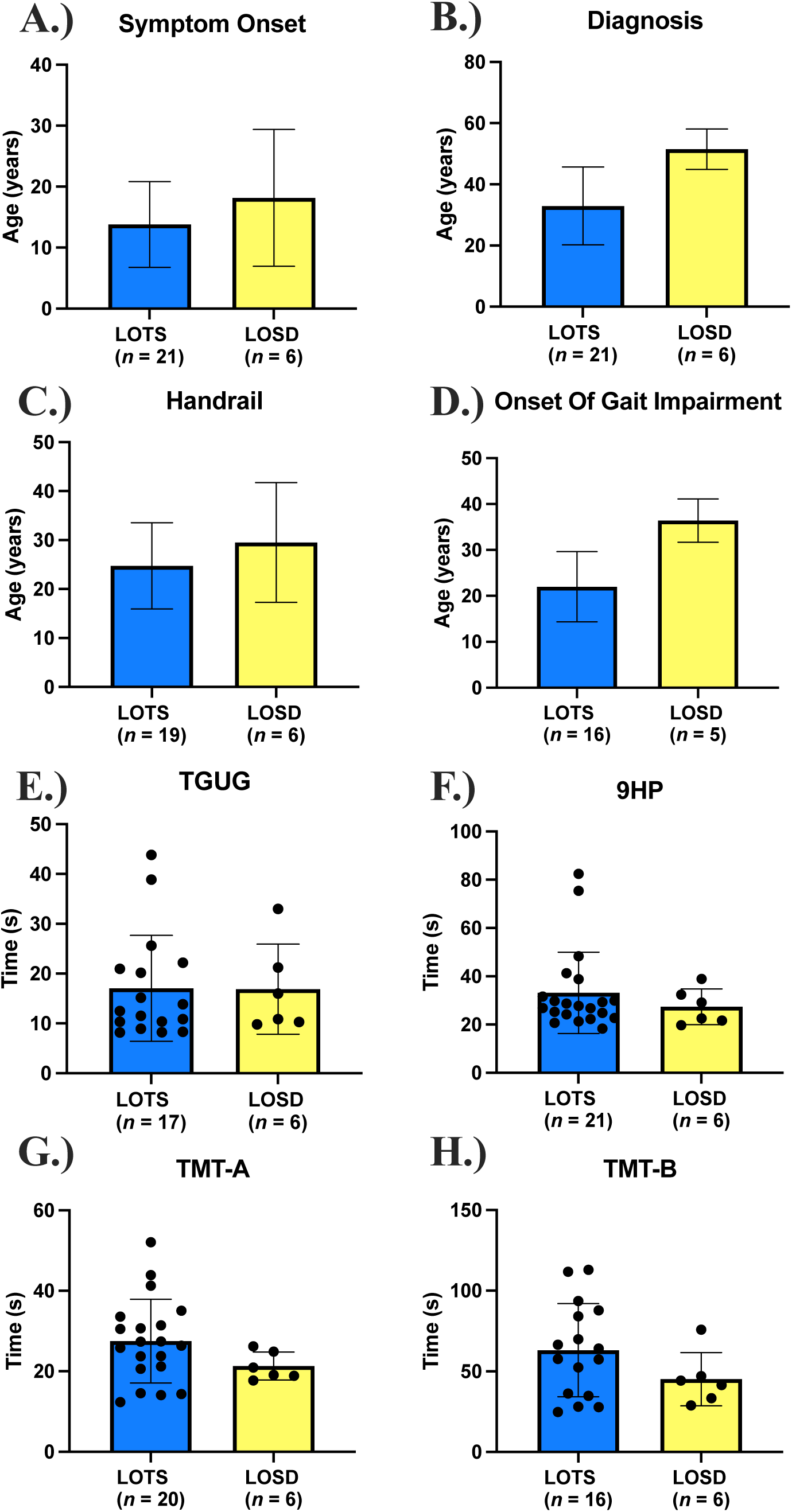
Clinical Phenotype and Clinical Outcome Assessments. **A.)** Age of Symptom onset between LOTS and LOSD (participant reported). **B.)** Age at diagnosis between LOTS and LOSD. **C.)** Age when Handrails were required. **D.)** Age at the onset of gait impairment. **E.)** Time get up and go (TGUG) functional testing. **F.)** 9-Hole Peg (9HP) Test on the dominant hand. **G.)** Trail Making Test (TMT) Part A. **H.)** Trail Making Test Part B. Individual data points were removed for Figure 1A-1D per MedRxiv requirements.

### Genotype

Fifteen of the 21 LOTS participants (71.4%) had one allele with the c.805G>A (p.G269S) variant, the most common variant associated with LOTS. An additional 3 participants (15.3%) were homozygous for this mutation.

Likely causative variants were identified in *HEXA* and *HEXB* in all individuals enrolled in the study and classified according to the 2015 Richards ACMG guidelines^56^ and the updated recommendations from the ACMG Sequence Variant Interpretation Working Group.^57–61^ Most participants were compound heterozygous for pathogenic (PATH) or likely pathogenic (LPATH) variants. In total, 16 variants were identified and classified; 9 in *HEXA* and 7 in *HEXB*; 13 were classified as PATH and 3 as LPATH (see Supplemental Table 1). These classifications align well with previous ClinVar entries, with the addition of one previously unseen LPATH variant as well as one large multi-exon deletion. Two participants (brothers) with the same nonsense HEXB variant did not have a variant in trans. Numerous functional studies support decreased activity in all 5 of the missense variants classified.^62–71^

### Functional Testing

TGUG (Figure 1E) were similar in LOTS (17.05 ± 10.64 seconds) and LOSD (16.87 ± 9.042 seconds). However, these values were qualitatively higher than the normative; healthy adults between 20–59 years achieve mean TGUG scores ranging from 8.57–9.90 seconds.^72^ For the 9HP test (Figure 1F) on the dominant hand, LOTS participants had slightly higher values on average with greater variability (33.18 ± 16.82 seconds) compared to LOSD participants (27.41 ± 7.412 seconds). Comparatively, normative data for the 9HP test shows 20–74-year-olds ranging between 16.1–22.0 seconds on the dominant hand.^73^ LOTS participants also tended to score higher on TMT-A (27.52 ± 10.41 seconds) than LOSD participants (21.31 ± 3.486 seconds, Figure 1G). These values generally fall within normative data, which reports the mean scores for TMT-A in 18–74-year-olds as ranging between 22.93 ± 6.87 seconds and 40.13 ± 14.48 seconds.^74^ Results for TMT-B (Figure 1H) were also higher in LOTS participants (63.14 ± 28.90 seconds) than LOSD participants (45.14 ± 16.52 seconds). Here, too, the LOTS and LOSD data fall around normative values, with mean values for TMT-B in 18–74-year-olds ranging between 48.97 ± 12.69 seconds and 86.27 ± 24.07 seconds.^74^

### Ataxia

Total BARS (out of 30, Figure 2A) were similar in LOTS (12.8 [IQR: 6.6, 16.75]) compared to LOSD (11.0 [IQR: 2.8, 14.6]). BARS finger-to-nose scores (out of 8, Figure 2F) were higher in LOTS (2.0 [IQR: 1.0, 2.0]) compared to LOSD (1.5 [IQR: 0, 2.0]), which was skewed as one LOTS participant with severe dysmetria scored an 8. BARS oculomotor scores (out of 2, Figure 2C) were higher in LOTS (0.25 [IQR: 0, 1.0]), compared to LOSD as all 6 LOSD individuals scored a 0, implying normal eye movement. BARS dysarthria scores (out of 4, Figure 2D) were higher in LOTS (0.5 [IQR: 0, 1.8]), compared to LOSD as all 6 participants with LOSD scored a 0 (normal speech). BARS knee-tibia scores (out of 8, Figure 2E) were similar between LOTS (8.0 [IQR: 2.0, 8.0]), and LOSD (8.0 [IQR: 1.5, 8.0]). BARS gait scores (Figure 2B) were also similar between LOTS (2.0 [IQR: 1.0, 6.0]) and LOSD (2.3 [IQR: 0.8, 5.0]). BARS knee-tibia scores and gait are substantially impacted by the associated severe lower extremity weakness in participants, precluding completion of the task that did not necessarily indicate ataxia.

**Figure 2.**
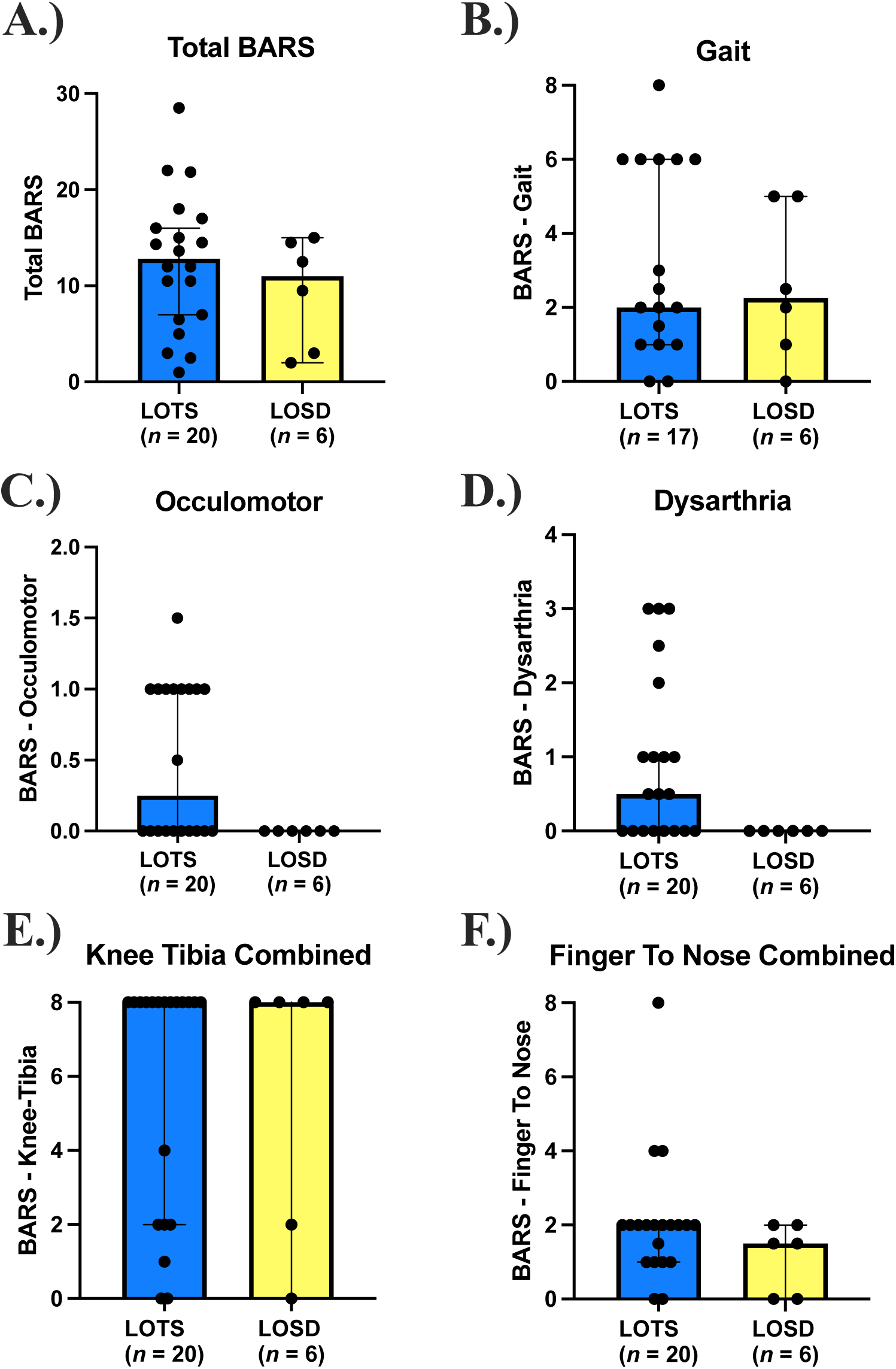
Brief Ataxia Rating Scale (BARS) Scores. LOTS participants are shown in blue and LOSD participants are shown in yellow. A. Total BARS score out of 30. B. BARS Gait Subdomain. C. BARS Oculomotor Subdomain. D. BARS Dysarthria Subdomain. E. BARS Knee Tibia Test (total of bilateral measurements). F. BARS Finger-to-nose test (total of bilateral measurements). Total BARS scores were scaled to 30 for individuals who were unable to ambulate and complete BARS Gait testing.

### Gait Analysis

Fourteen LOTS (8 female) and 4 LOSD (2 female) participants were tested alongside eighteen age- and sex-matched controls (one control per participant) from MacKay et al.^37^ As shown, abnormalities were observed on all gait parameters (Figure 3A-3D). As a group, LOTS and LOSD participants (88.73 ± 36.49 cm/s and 97.31 ± 33.24 cm/s, respectively) walked slower than controls (130.34 ± 5.87 cm/s). LOTS participants had a shorter step length than controls (56.02 ± 14.83 cm vs. 68.03 ± 2.82 cm), but the difference was less prominent in the LOSD cohort (61.34 ± 10.24 cm vs. 68.03 ± 2.82 cm). Additionally, both LOTS and LOSD participants had a wider bases of support as evident by step width (14.29 ± 5.27 cm and 11.11 ± 1.36 cm respectively) than controls (8.18 ± 0.65 cm), and both spent longer time in the double support phase of gait (LOTS 0.52 ± 0.25 s, LOSD 0.49 ± 0.19 s, controls 0.24 ± 0.02 s). Individual values vary, with a few participants scoring within the normal range.

**Figure 3.**
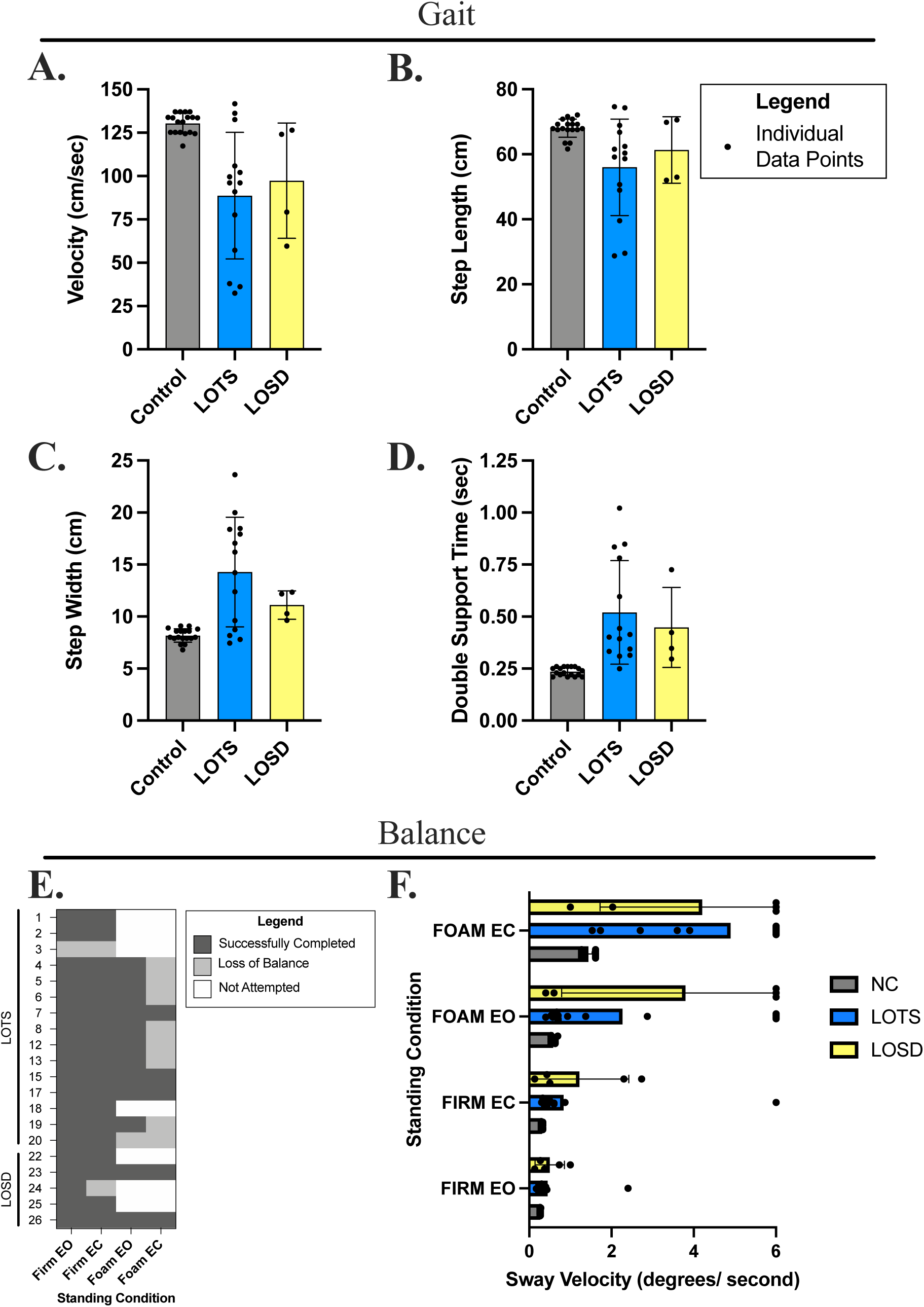
Gait and balance testing results. Gait parameters recorded for LOTS (blue), LOSD (yellow), and their age/sex matched controls (grey). **A.)** Gait velocity (cm/s), **B.)** step length (cm), **C.)** step width (cm), and **D.)** double support time (sec) are shown for each group. Data are presented as individual data points with the group mean ± SD, *n* = 15 LOTS (9 female, 6 male), *n* = 5 LOSD (2 female, 3 male), *n* = 18 control (10 female, 8 male). **E.)** mCTSIB Balance test completion per condition displayed by cohort (LOTS and LOSD) and participant number (Table 1). Participants who successfully completed all 3 trials for that condition are shown in dark gray, participants who lost balance during at least one trial are shown in light gray, and participants who did not attempt the testing are shown in white. **F.)** mCTSIB sway velocity of LOTS participants, LOSD participants, and their age/sex matched controls per condition in order of difficulty (top to bottom). Data are represented as individual data points with the group mean marked as a diamond (blue for LOTS, yellow for LOSD, and grey for controls). Note: Foam EC = standing on foam surface with eyes closed, Foam EO = standing on foam surface with eyes open, Firm EC = standing on firm surface with eyes closed, Firm EO = standing on firm surface with eyes open.

### Balance Testing

Most participants were able to stand a on firm surface with eyes open or closed without losing their balance (Figure 3E). However, only 12 participants (10 LOTS and 2 LOSD) were able to stand on the foam surface with their eyes open, and only 3 LOTS and 2 LOSD participants were able to successfully complete all trials of the test without losing their balance.

#### Sway Velocity - Firm Condition

Sway velocity on the firm surface with eyes open was similar between the three cohorts, although one severely impaired individual in the LOTS group stands out with a high score (Figure 3F). Average sway velocity was higher for the LOTS and LOSD cohorts than controls under the firm condition that required eyes closed, but the results are skewed by a few individuals.

#### Sway Velocity - Foam Condition

Sway velocity was higher in the foam condition with eyes open in both the LOTS and LOSD compared to controls, however this was skewed by the subset of individuals (5 LOTS and 3 LOSD) unable to successfully complete the testing. Similarly, while standing on the foam with eyes closed, LOTS and LOSD also had higher sway velocities compared to controls.

### Quadriceps and Hamstring Strength Testing

Eighteen LOTS, 5 LOSD, and 19 control participants completed the testing. There was no qualitative difference between left and right isokinetic torques or between left and right isometric torques in all three groups (Figure 4A and 4B). Isokinetic and isometric extension was qualitatively larger in the control group (left isokinetic: 1.5 ± 0.5 Nm/kg; left isometric: 2.0 ± 0.5 Nm/kg) than in the LOTS (left isokinetic: 0.25 ± 0.2 Nm/kg; left isometric: 0.25 ± 0.25 Nm/kg) and LOSD (left isokinetic: 0.25 ± 0.2 Nm/kg; left isometric: 0.25 ± 0.25 Nm/kg) groups. However, there was no qualitative difference in isokinetic and isometric flexion between the control (left isokinetic: 0.7 ± 0.3 Nm/kg; left isometric: 0.9 ± 0.5 Nm/kg), LOTS (left isokinetic: 0.4 ± 0.2 Nm/kg; left isometric: 0.7 ± 0.3 Nm/kg), and LOSD (left isokinetic: 0.6 ± 0.2 Nm/kg; left isometric: 0.7 ± 0.3 Nm/kg) groups. Control group isokinetic extension (∼1.5 Nm/kg) was approximately twice as large as isokinetic flexion (∼0.7 Nm/kg). Conversely, LOTS isokinetic extension (∼0.25 Nm/kg) was approximately half as large as isokinetic flexion (∼0.4 Nm/kg). The LOSD group had a similar pattern to the LOTS group.

**Figure 4.**
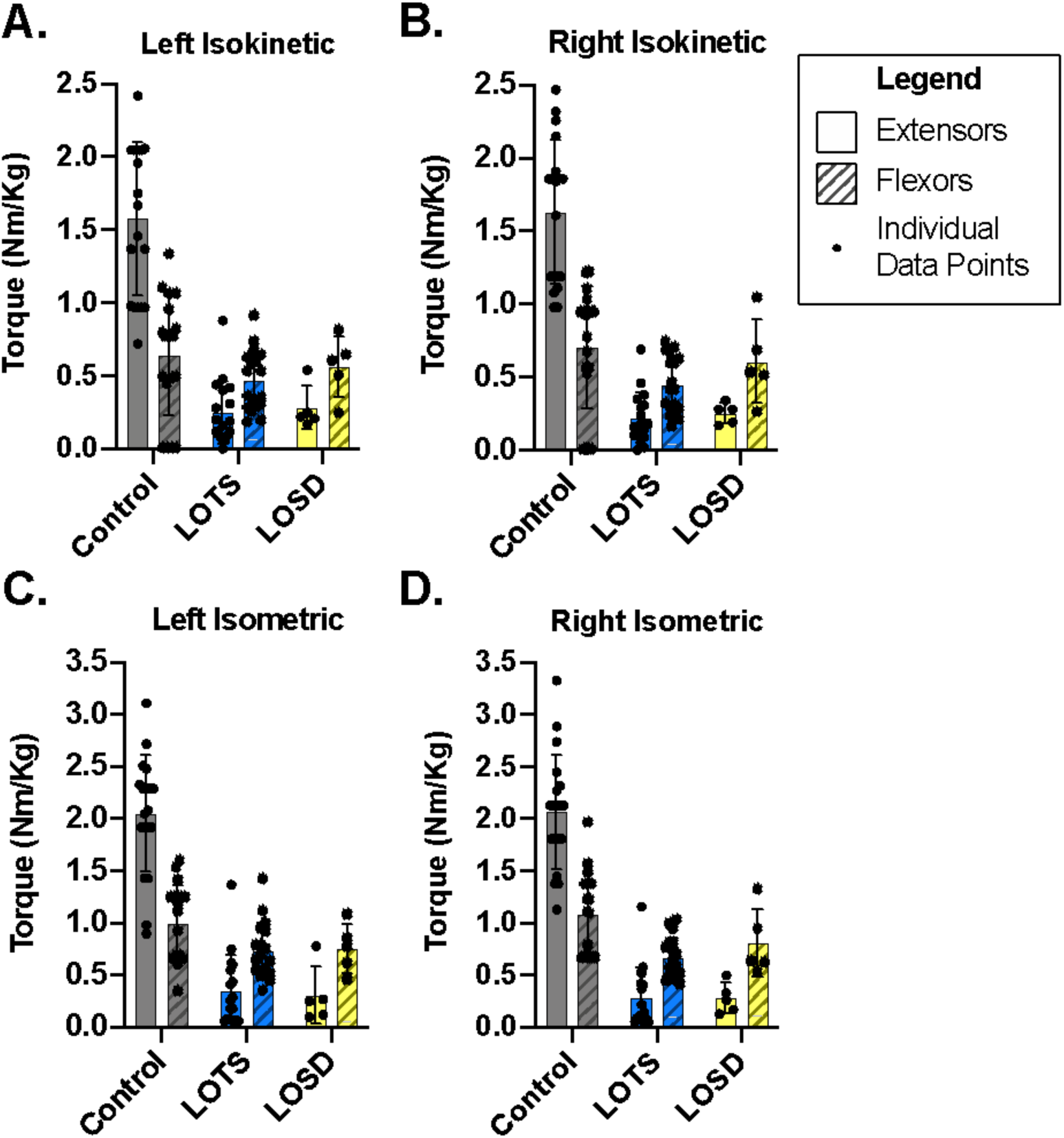
Knee extensors (solid fill) and flexors (striped fill) strength in LOTS (blue) and LOSD participants (shown in yellow), and their age/sex matched controls (shown in grey). A. Left isokinetic, B. right isokinetic, C. left isometric, and D. right isometric torque normalized to the participant’s weight (Nm/kg) are shown for each group. Data are presented as individual data points with the group mean ± SD, *n* = 18 LOTS (11 female, 7 male), *n* = 5 LOSD (2 female, 3 male), *n* = 19 control (10 female, 9 male). Age/sex matched controls were not available for 3 LOTS participants and 1 LOSD participant.

### Sensory Examination and Electrodiagnostic Testing

The sural and median sensory amplitudes were lower in the LOSD group (Figure 5). The motor amplitudes were not different between the two groups. Sural Sensory Nerve Action Potentials (SNAPs) were absent in 83% of LOSD (5/6). The remaining LOSD participants had an amplitude at the normal amplitude threshold (6 µV). This contrasts with 33% of LOTS (5/15) with absent sural nerve SNAPs, 2 (13%) with low amplitude SNAPs and 7 (45%) with normal sural SNAPs. Similarly, all LOSD participants had reduced (4/5), or absent 1/5 median nerve SNAP amplitude. The mean median SNAP amplitude in LOSD was 4.0µVcompared to normal laboratory control threshold of >= 15 µV), whereas, LOTS participants, median nerve SNAP amplitudes were normal in 9/15 participants (60%), absent in one (6%) and below normal amplitude in 5/15 (33%). The mean median nerve SNAP amplitude in LOTS was 23.7µV. The motor nerve conduction studies show less abnormalities compared to the sensory nerve conduction studies. Compound Motor Action Potential (CMAP) amplitudes in the tibialis anterior muscle were low in 3/15 (20%) participants with LOTS and in 1/5 with LOSD (20%). Median nerve CMAP amplitudes were normal in both participant group.

**Figure 5.**
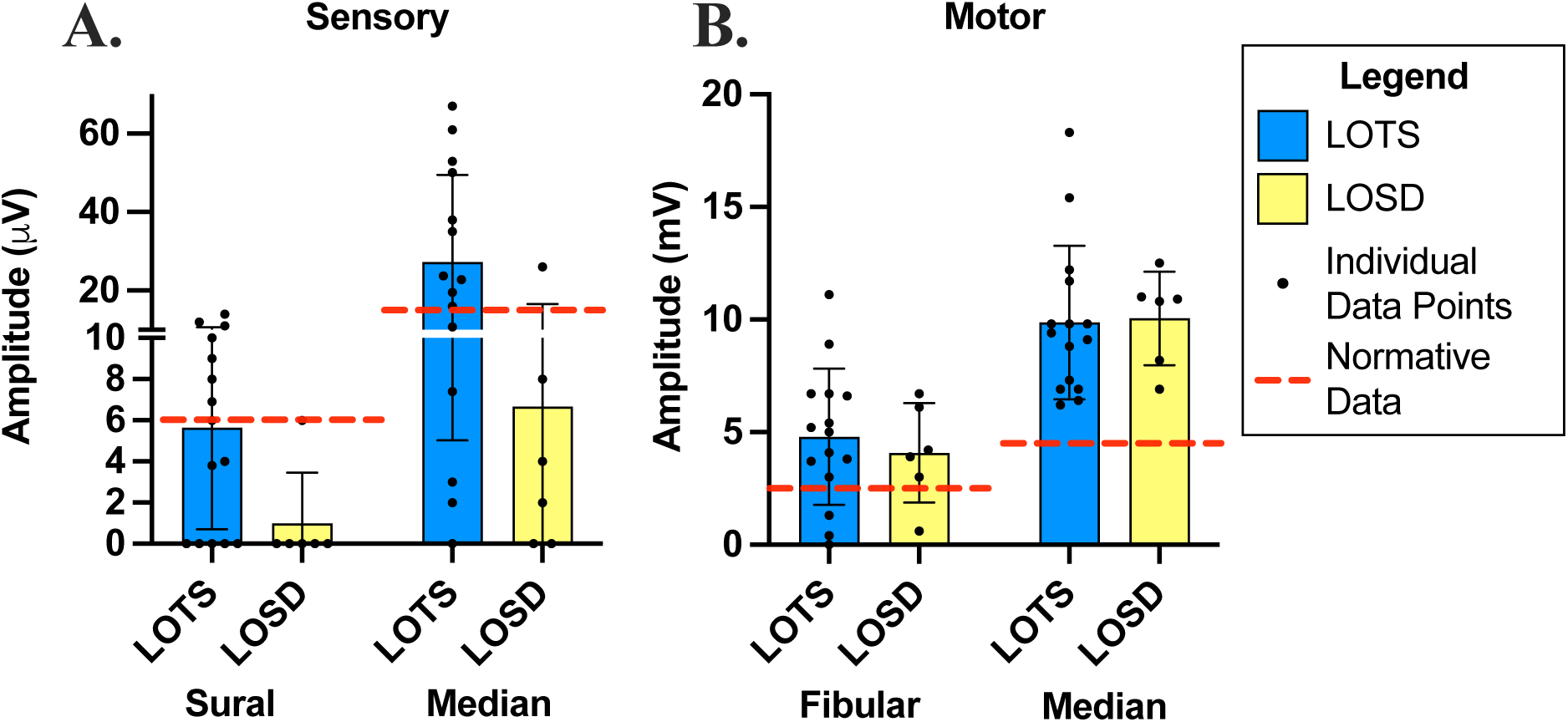
Electrodiagnostic amplitudes of A. sensory nerves (SN) and B. motor nerves (MN) recorded for LOTS and LOSD participants. LOTS participants, *n* = 15 (10 female, 5 male), are shown in blue and LOSD participants, *n* = 6 (3 women, 3 male) are shown in yellow. [Normal values: sural SN ≥ 6.0 µV, median SN ≥ 15.0 µV, fibular MN ≥ 2.5 mV, median MN ≥ 4.5 mV (shown in red dashed line).]

Both LOTS and LOSD participants had acute and chronic neurogenic changes on needle EMG (fibrillations, positive sharp wave and or complex repetitive discharges) on needle EMG evaluation in the triceps, quadriceps, medial gastrocnemius, and tibialis anterior muscles though slightly more prevalent in the LOTS participants. Overall, the needle EMG findings were not length-dependent and more consistent with motor neuronopathy and, as expected, with more severe involvement of motor neuron corresponding to the myotomes with more severe muscle weakness.

### MRI Analysis

18 LOTS and 6 LOSD participants had MRI testing at the cross-sectional evaluation. Gray matter volume (GMV) was similar between NC (46.8 ± 7.3 percent of ICV), LOTS (47.2 ± 6.1 percent of ICV), and LOSD (46.6 ± 3.4 percent of ICV). White matter volume (WMV) was similar between NC (33.3 ± 6.0 percent of ICV), LOTS (33.6 ± 4.1 percent of ICV), and LOSD (36.3 ± 2.6 percent of ICV). Cerebellar volumes were lower in LOTS (7.6 ± 2.3 percent of ICV) compared to both NC (10.2 ± 1.8 percent of ICV) and LOSD (10.5 ± 1.0 percent of ICV). 4^th^ ventricle volume was higher in LOTS (0.20 ± 0.06 percent of ICV) compared to both NC (0.13 ± 0.05 percent of ICV) and LOSD (0.12 ± 0.03 percent of ICV). When analyzed at the lobule level, LOTS participants showed decreased volumes of both lobule V and VI (0.29 ± 0.06, 0.62 ± 0.24) compared to LOSD (0.34 ± 0.03, 0.84 ± 0.04) and NC (0.37 ± 0.05, 0.95 ± 0.14), respectively.

## Discussion

Despite shared biochemical deficiencies of ß-hexosaminidase A and significant clinical overlap, differences have emerged between late-onset Tay-Sachs (LOTS) and Sandhoff diseases (LOSD). This study aimed to further characterize this dichotomy through the cross-sectional phenotypic evaluations of 21 participants with LOTS and 6 participants with LOSD. We found both LOTS and LOSD to frequently have lower limb weakness later progressing to upper limb weakness, dysphagia, and tremor (Figure 7). LOTS participants predominately had signs and symptoms of cerebellar dysfunction including oculomotor symptoms, psychiatric symptoms, cerebellar ataxia, and dysarthria. Whereas LOSD participants had length dependent sensory neuropathy which was not present in LOTS.

In a natural history study by Masingue et al. describing 57 adult participants with GM2 gangliosidosis, they found three main clinical presentations: lower limb weakness progressing to upper limb weakness, cerebellar ataxia, psychiatric symptoms (only in LOTS), or some combination of the three.^17^ The 26 GM2 gangliosidosis participants described in this study are similar, as the probands all fit into one of the ascribed categories. Lower limb weakness was highly prevalent in both LOTS and LOSD as evidenced by gait disturbances, strength testing, balance testing, presenting symptom(s), and the high frequency of participants requiring handrails to climb stairs. Upper limb weakness primarily affecting the triceps muscles was also highly prevalent in both LOTS and LOSD, occurring later in the disease course.

Isokinetic and isometric strength of both the hamstrings and quadriceps was found to be impaired in both groups of participants, with the knee extensors being profoundly weak. As a group, average knee extension strength was roughly 10% of predicted when compared to age and sex matched controls. These results confirm our clinical observations, and reports in the literature, of knee extensors being more affected by both disease processes than knee flexor muscle groups.^10,17^ The rectus femoris, a bi-articular muscle acts both as a knee extensor and as a hip flexor. Therefore, the observed hip flexor weakness in LOTS and LOSD is likely to be directly related, at least in part, to the observed profound atrophy and weakness in the rectus femoris muscle. Lower limb muscle strength including hip flexion and knee extension are crucial components for assuming an upright position, maintaining an upright stance and controlling gait and balance, all of which were altered in LOTS and LOSD compared to normative controls.

Ataxia, as assessed by BARS, showed similarities between LOTS and LOSD including lower extremity scores for dysmetria, as most participants were unable to properly execute the testing procedure due to lack of proximal antigravity strength to complete the task. We encountered difficulties parsing the exact contributions of different neurological deficits to the overall experience of imbalance and falls in our study population, especially weakness and cerebellar deficits in LOTS and weakness and proprioceptive deficits with sensory ataxia in LOSD. As strength plays a major role on one’s ability to complete balance testing, we frequently observed participants in both cohorts locked their knees to maintain upright postural control, counterbalancing the quadriceps weakness during the mCTSIB balance test. In fact, many participants reported they could not rely on knee strength and were afraid if their knees buckled, they would collapse and could not prevent falling.

However, the BARS highlighted two considerable disparities between LOTS and LOSD. The oculomotor portion of the BARS was abnormal in LOTS and normal to LOSD. Saccadic dysmetria has previously been described in LOTS,^10,75^ and further described in a cohort including 7 LOTS participants and 3 LOSD participants.^76^ Our study further validates the finding that oculomotor dysfunction is prevalent in LOTS than LOSD. It is well known that cerebellar dysfunction affects the oculomotor system in many ways including saccadic intrusion, “square wave” jerks, saccadic dysmetria, abnormal VOR and disruption of normal saccade velocity.^10^ Cerebellar lobule topologies in relation to oculomotor abnormalities localizes primarily to lobules V, VI and VII, and both Crus I and II.^77,78^

Furthermore, speech articulation was also substantially altered in most (71%) LOTS participants compared to LOSD and in none of the participants with LOSD. Progressively worsening dysarthric speech has been demonstrated in late-onset GM2 gangliosidosis,^79^ and isolated dysarthria has even been described as an initial clinical presentation in LOTS.^21^ This study diverges from previous reports showing an absence of dysarthria in our LOSD participants while illustrating a wide spectrum of variability in LOTS from, participants with long disease course and entirely normal speech articulation to younger participants with profound intelligibility challenges. LOTS-associated dysarthria is characterized by distinct and complex features including abnormal articulation, explosive features and rapid pace among others. Dysarthria in LOTS has been attributed to cerebellar dysfunction, and indeed, there is supporting evidence that dysfunction or injury to lobules V and VI are linked to dysarthric speech in other settings.^80^

Neuropsychiatric symptoms including the emergence of acute mania and psychosis, even as a presenting symptom, have long been known in LOTS.^81,82,83^ Indeed, in this cross-sectional evaluation, 5/21 (24%) of the LOTS participants and none of the LOSD participants had a history of psychiatric symptoms, and 2/16 (13%) of the LOTS probands in this study had psychosis as the presenting symptom. Our observation is consistent with Masingue et al.,^17^ where in a combined cohort of 57 participants, including participants evaluated at their center and participants reported from other studies (40 LOTS and 17 LOSD), psychiatric symptoms were the initial presentations of LOTS for 11/40 (28%) participants, while none of the 17 LOSD participants presented with psychiatric symptoms. Only 1 LOSD participant developed psychiatric symptoms later in the disease course.^17^

The basis for neuropsychiatric symptoms in LOTS and the apparent lower prevalence in LOSD is not fully understood, however, the finding of more profound cerebellar atrophy in LOTS vs. LOSD may provide a clue to these differences. There is growing evidence demonstrating that cerebellar dysfunction, and more specifically the posterior cerebellum, has an impact on behavior. The cerebellar cognitive affective syndrome (CCAS), encompassing several cognitive and behavioral deficits, is caused by damage to the posterior cerebellum (lobules VI, VII, and IX), and results in impaired executive function and affect regulation.^84,85,86^

Magnetic resonance imaging (MRI) demonstrated expected results based on previous reports.^26,27,29,55^ First, both supratentorial gray and white matter volumes were not influenced by LOTS or LOSD (Figure 6). LOTS participants had reduced cerebellar volume (including lobules V and VI) and associated enlargement of the 4^th^ ventricle. While the differential finding of cerebellar atrophy in LOTS vs. LOSD has been described in prior scoping review,^28^ this finding, including its detailed the cerebellar lobule topology, has not been extensively quantified volumetrically until recently.^55^ The discrepancy in both cerebellar atrophy and psychiatric and speech manifestations in LOTS versus LOSD imply that severe cerebellar atrophy and perhaps, even cerebellar hypoplasia, contribute to both these clinical findings.

**Figure 6.**
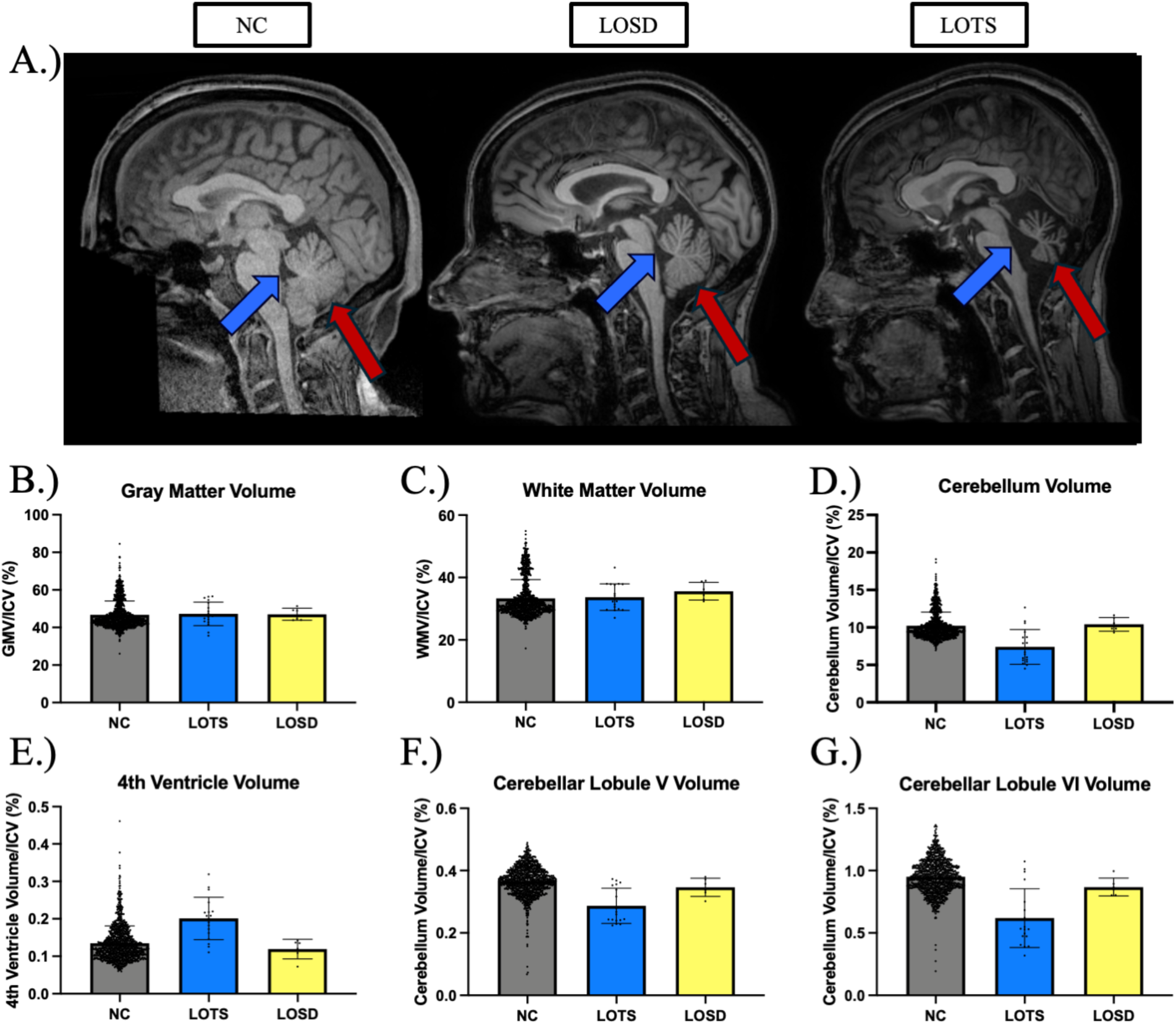
MRI Analysis. **(A)** Sagittal slice of one 41-45 year-old female neurotypical control (NC), one 41–45-year-old late-onset Sandhoff disease (LOSD) participant, and one 41–45-year-old late-onset Tay-Sachs (LOTS) participant. The blue arrows designate the 4^th^ ventricle, and the red arrows designate the cerebellum of the three participants. The LOTS participant demonstrates considerable 4^th^ ventricle enlargement and cerebellar atrophy compared to both the LOSD participant and NC. **(B)** Volumetric analysis of gray matter volume (GMV) between NC, LOTS, and LOSD participants. **(C)** Volumetric analysis of white matter volume (WMV) between NC, LOTS, and LOSD participants. **(D)** Volumetric analysis of cerebellum volume between NC, LOTS, and LOSD participants. **(E)** Volumetric analysis of 4^th^ ventricle volume between NC, LOTS, and LOSD participants. **(F)** Volumetric analysis of cerebellar lobule V and **(G)** lobule VI between LOTS and LOSD. All volumes were controlled for total intracranial volume (ICV). LOTS participants are shown in blue (*n* = 18), LOSD participants are shown in yellow (*n* = 6), and neurotypical controls are shown in gray (*n* = 1038).

**Figure 7.**
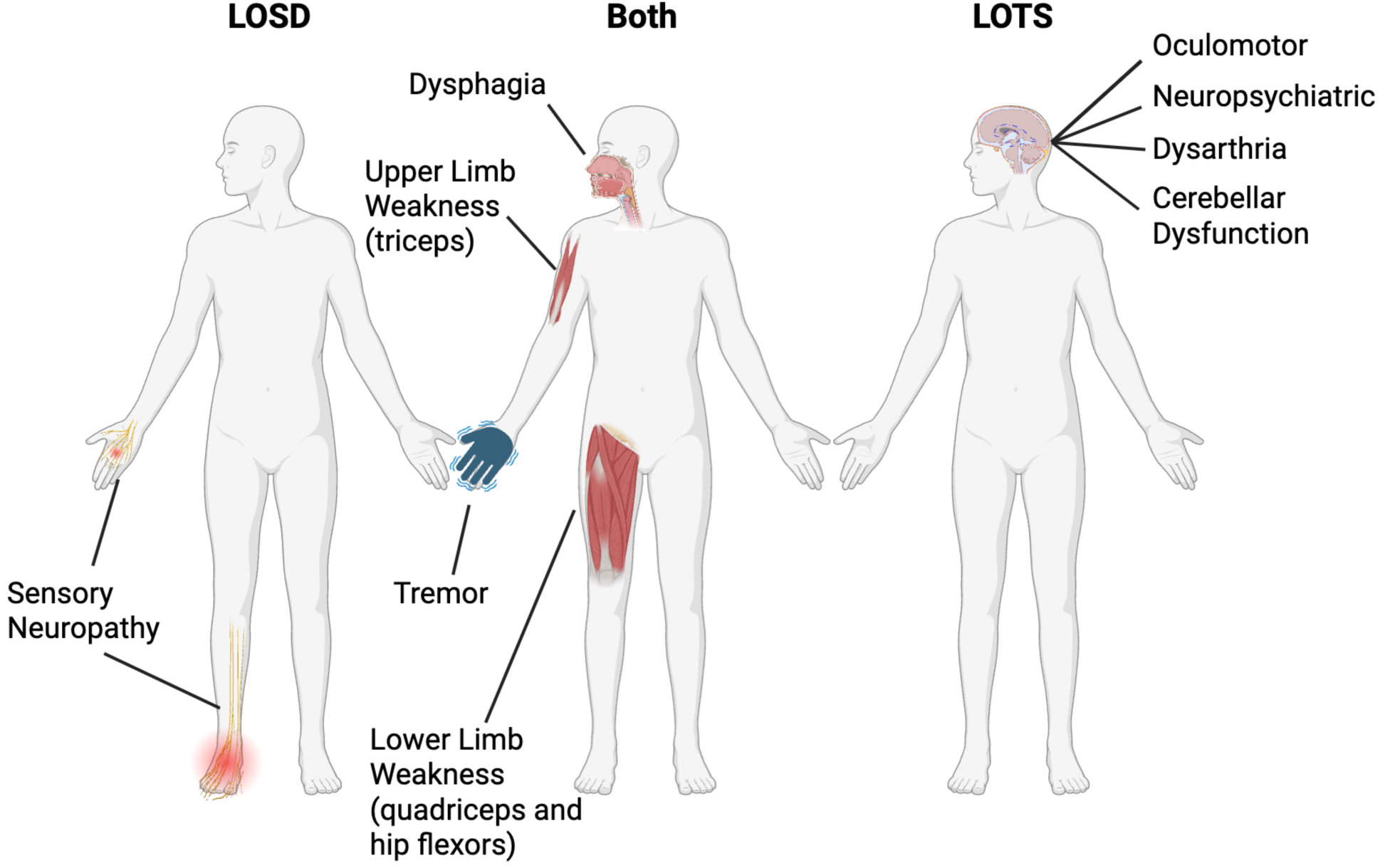
Illustration of symptom similarities and differences between late-onset GM2 gangliosidoses (both), late-onset Tay-Sachs (LOTS), and late-onset Sandhoff disease (LOSD).

Analysis of the individual cerebellar lobules has been described in LOTS,^29,55^ demonstrating atrophy in the posterior cerebellum. However, the study by Májovská et al.,^29^ found no relationship between cerebellar lobule atrophy and presence of psychiatric symptoms. However, their analysis included only seven participants with psychiatric impairment, and the participants they include in their analysis a heterogeneous group of psychiatric symptoms including depression, anxiety, adjustment disorder, and cognitive deficits.^29^ This may indicate the relationship neuropsychiatric symptoms and cerebellar atrophy is not direct, and multiple cerebellar associated clinical features (dysarthria, oculomotor dysfunction, psychiatric impairment, and cerebellar ataxia) should be taken in consideration with cerebellar atrophy. More detailed phenotyping including detailed neuropsychiatric analysis and MRI-derived cerebellar lobule analysis including cerebellar white matter diffusion tensor imaging is needed.^27^

Clinical and electrodiagnostic studies in our cohort present an additional element of the LOTS versus LOSD phenotypic dichotomy. While both LOTS and LOSD participants appear to experience identical patterns and severity of lower motor neuronopathy with predilection for hip flexors, knee extensors and triceps muscles, LOSD but not LOTS participants in our cohort showed substantial and disabling sensory deficits affecting all sensory modalities (proprioception, vibration, temperature and pinprick), and reflected in symptoms of dysautonomia including and not limited to reported symptoms of chronic diarrhea, thermoregulatory deficits, and heat-related acro-paresthesia, present in all of our LOSD participants. Severe proprioceptive deficits in LOSD loss further contribute to their sense of imbalance. While modest sensory NCV deficits have been reported in a prior LOTS study, LOTS participants are seldom symptomatic.^17^

This study, although one of the largest reported single cohorts of late-onset GM2 participants, is principally limited by the sample size (*n* = 27). This highlights the rarity of the disease and likely both its under- and delayed diagnosis. This study is also limited by incomplete data collection due to personal choice or safety concerns. Furthermore, we scored ataxia using the BARS scale by a single rater, therefore, interrater reliability was not established. The late-onset GM2 gangliosidoses are much more than an ataxic syndrome. Further efforts should be undertaken to develop disease specific scales that include neuromuscular weakness, dysarthria, cerebellar motor and cognitive deficits (CCAS), and sensory dysfunction. Participant- and caregiver-related outcomes in relation to disease experience should be incorporated as a part of this effort.^30^ The normative controls for MRI analysis were scanned using different scanners and acquisition protocols, a limitation of the MRI results, however all the acquisition protocols were of high resolution (1 mm^3^ isotropic voxels or better), similar to the study participants.

In conclusion, this study highlights key differences between LOTS and LOSD including cerebellar dysfunction, imaging abnormalities, and sensory neuropathy. LOTS participants showed increased prevalence and severity of dysarthric speech, oculomotor abnormalities, and neuropsychiatric symptoms, all of which can relate to cerebellar dysfunction. In contrast, LOSD participants experienced sensory deficits, not present in the participants with LOTS. Future studies are needed to understand the genetic and molecular explanation for the differences between LOTS and LOSD despite a shared enzymatic defect.

## Data availability

The GM2 data described in this manuscript are available from the corresponding author upon reasonable request. Neurotypical control MRI data are available from OpenNeuro at the following links; NIMH: https://doi.org/10.18112/openneuro.ds004215.v1.0.0,45,46 Neurocognitive Aging Data Release with Behavioral, Structural, and Multi-echo functional MRI measures: https://openneuro.org/datasets/ds003592/versions/1.0.13,^47,48^ Paingen_Placebo: https://openneuro.org/datasets/ds004746/versions/1.0.1,^49,50^ AgeRisk: https://openneuro.org/datasets/ds004711/versions/1.0.0.^51,52^

## Acknowledgements

We thank the participants and their families for the generosity of their time and efforts. We are also grateful to many staff members at the NIH Clinical Center to the participants’ local care providers who have contributed their expertise over many years. Magnetic resonance imaging analysis in this work utilized the computational resources of the Biowulf Linux cluster at the National Institutes of Health (http://hpc.nih.gov).

## Ethics Declaration

The NIH Institutional Review Board approved this protocol (02-HG-0107). Informed consent was completed with all participants prior to participation, and all research was completed in accordance with the Declaration of Helsinki.

## Supplementary Materials

**Supplement Figure 1.**
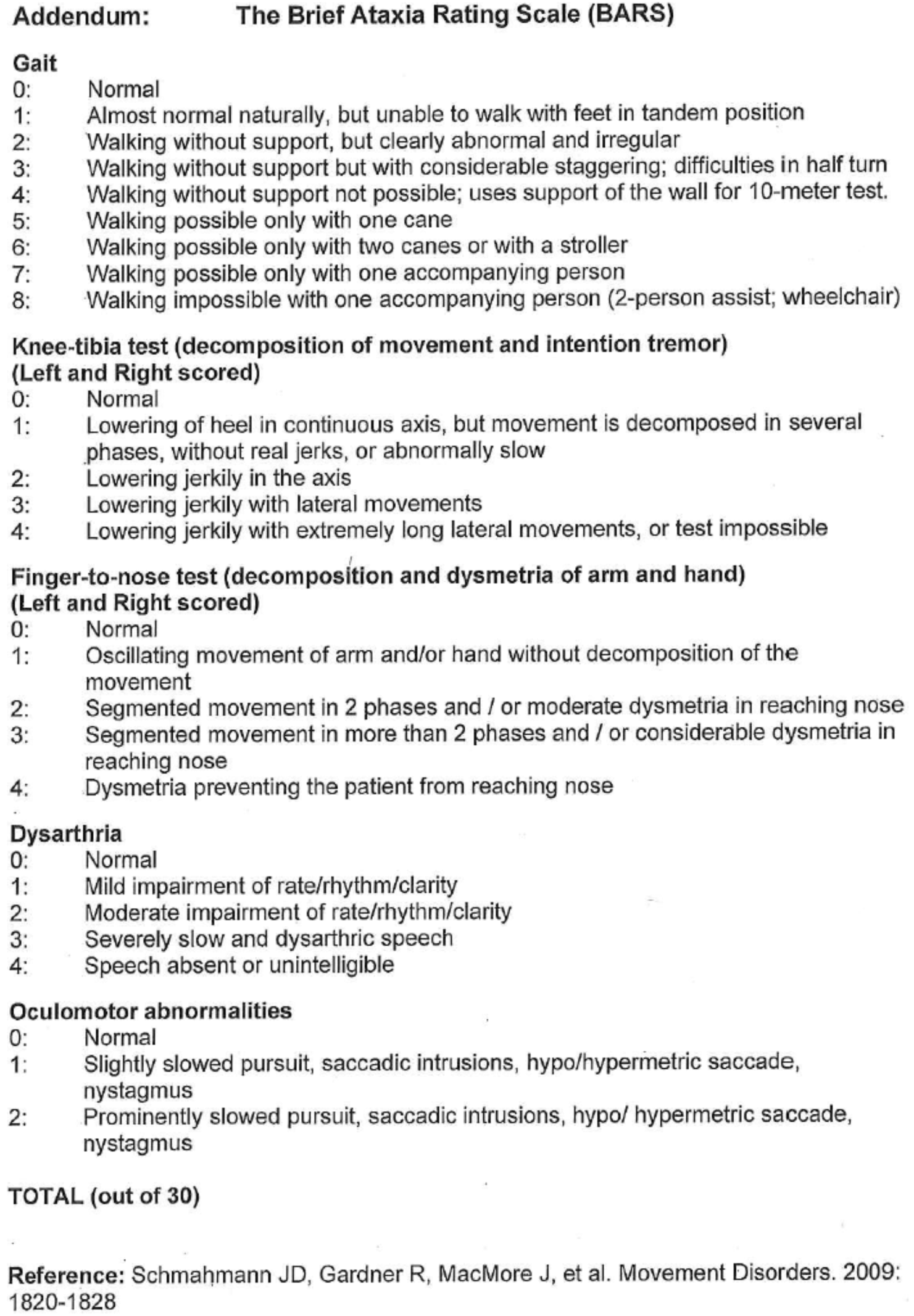
The Brief Ataxia Rating Scale (BARS).

